# How often do symptoms return after unsuccessful drug treatment for malaria? A systematic review and meta-analysis

**DOI:** 10.1101/2020.08.21.20179382

**Authors:** Rida Mumtaz, Lucy C. Okell, Joseph D. Challenger

## Abstract

**Background:** In clinical trials of therapies for uncomplicated *Plasmodium falciparum*, there are usually some patients who fail treatment even in the absence of drug resistance. Treatment failures are categorised as ‘clinical’ or ‘parasitological’ failures, the latter indicating that recrudescence of the infection has occurred without inducing the return of symptoms. Asymptomatic treatment failure has public health implications for continued malaria transmission and may be important for the spread of drug-resistant malaria. As the number of treatment failures in an individual trial is often low, it is difficult to assess how commonplace asymptomatic treatment failure is, and with what factors it is associated.

**Methods:** A systematic literature review was carried out on clinical trials of artemether-lumefantrine (AL) in patients seeking treatment for symptomatic uncomplicated falciparum malaria, and information on symptoms during treatment failure was recorded. Only treatment failures examined by polymerase chain reaction (PCR) were included, so as to exclude reinfections. Using a multivariable Bayesian regression model, we explored factors potentially explaining the proportion of recrudescent infections which are symptomatic across the trials included in our study.

**Results:** Across 60 published trials including 9137 malaria patients we found that 40.8% (95% CIs [35.9-45.8%]) of late treatment failures were symptomatic. We found a positive association between transmission intensity and the observed proportion of treatment failures that were asymptomatic. We also found that symptoms were more likely to return in trials that only enrolled children aged < 72 months (odds ratio =1.62, 95% CIs [1.01,2.59]). However, 84 studies had to be excluded from our analysis, as treatment failures were not specified as symptomatic or asymptomatic.

**Conclusions:** AL, the most widely used treatment for uncomplicated *Plasmodium falciparum* in Africa, remains a highly efficacious drug in most endemic countries. However in the small proportion of patients where AL does not clear parasitaemia, the majority of patients do not develop symptoms again and thus would be unlikely to seek another course of treatment. This continued asymptomatic parasite carriage in patients who have been treated may have implications for drug-resistant parasites being introduced into high-transmissions settings.

## Background

Artemisinin combination therapies (ACTs), the WHO-recommended drugs for uncomplicated *Plasmodium falciparum*, remain highly effective in Africa (1). All require a three-day dosing regimen, with either one or two doses a day (2). However, even if dosing and patient adherence to treatment is optimal, as occurs in clinical trials with directly observed treatment, the treatment will fail to fully clear parasites in a small proportion of patients, usually under 5% (3). Pharmacokinetic studies of drug treatment demonstrate that the inter-individual variation in the absorption and elimination of drugs is very large, and has been linked to treatment outcome in a number of studies (4–8). There is evidence that severe acute malnutrition can inhibit drug absorption (9), and that pharmacogenetic factors play a role (10,11). In routine healthcare settings, additional factors can cause treatment to fail, such as imperfect adherence to the dosing regimen (12,13). Some frontline drugs, such as lumefantrine, require that doses be taken with fatty food or drink to ensure optimal absorption (14).

Treatment failures have a number of undesirable consequences: most immediately for the patient concerned, who may feel better initially but become symptomatic again as the parasites which have survived treatment multiply once more. In addition, the patient may be able to infect a feeding mosquito thus contributing to malaria transmission. Furthermore, a parasite population being exposed to, but not completely cleared by, antimalarial drugs has implications for the development of drug resistance. During the evolution and spread of drug resistance, drug-sensitive parasites coexist with more resistant parasites. Some patients have genetically distinct subpopulations of parasites within their infection, either because mutations have occurred *de novo* as the parasites multiplied during the current infection, or because mosquitoes have injected more than one parasite clone in a single or multiple bites. If there are any parasites with some degree of resistance to the drug within a patient’s infection, they have a survival advantage over more sensitive parasites. Those which survive treatment are more likely to be transmitted to others. In this regard, an important consideration is whether patients who have failed treatment become symptomatic again. The reoccurrence of symptoms, although unpleasant for patients, may have positive implications at the community level. If a patient’s recrudescent infection is treated again, the capacity for further transmission is diminished (15). Even if parasites are partially resistant, the treatment has a chance of being successful the second time, since very few types of drug resistance confer 100% lack of susceptibility to drugs. Although patients who do not become symptomatic after the infection recrudesces may be identified and retreated in a clinical trial, this is very unlikely in routine healthcare settings. Untreated infections of falciparum malaria can be very long lasting (16,17), during which time it may be possible to infect feeding mosquitoes.

One can gain insight into the proportion of treatment failures that become symptomatic from clinical trials. Typically, treatment failures that are recorded in a trial are stratified into different categories. Early treatment failures (ETFs) indicate that the patient did not respond to treatment, and fall into one of four categories: (i) Indications of severe malaria on day 1, 2, or 3 in a patient who is still parasite positive; (ii) Parasitaemia on day 2 that is higher than observed on admission; (iii) Parasitaemia on day 3 with axilliary temperature ≥ 37.5; (iv) Parasitaemia on day 3 ≥ 25% of that observed on admission (18). Late treatment failures (LTFs) indicate that although patients responded to treatment and clinical symptoms abated, the drug treatment did not completely clear the parasite population, although the parasite density may become too low to be detectable by microscopy. If the infection is not cleared before drug concentrations decline below effective concentrations, the parasite population can grow in size again. For ACTs, late treatment failures are rarely observed in the first two weeks (see e.g. Ref. (19)), and patients in clinical trials should be followed up for at least 28 days (18). Late parasitological failures (LPFs) indicate that patients were found to be parasite-positive during follow-up without the presence of symptoms (axilliary temperature < 37.5°C). Late clinical failures (LCFs) indicate that patients were symptomatic (indications of severe malaria or an axilliary temperature ≥ 37.5°C) and positive for parasites (18). Infections detected during the follow-up period of a trial should be evaluated by polymerase chain reaction (PCR), to discriminate between a recrudescent infection and an infection acquired subsequently. In an individual trial, the number of PCR-corrected treatment failures is often very small (3), which means that it is hard to gain insight into the likelihood of symptoms returning after parasitaemia rebounds.

Here, we conduct a systematic review of clinical trials to quantify the proportion of late treatment failures that were recorded as being symptomatic failures (LCFs). Intuitively, this proportion will depend on the drug treatment administered: drugs that are eliminated more slowly from the body should suppress parasite densities for a longer period of time. Therefore, we restrict ourselves to clinical trials of a single therapy, the ACT artemether-lumefantrine (AL). As of 2017, AL is a first or second line treatment for uncomplicated *P. falciparum* malaria in 59 countries across the world, including 32 in Africa (20). Specifically, we searched for trials that stratified late treatment failures into LCFs and LPFs after PCR-correction was carried out. Our literature review built upon a Cochrane review of ACT treatments (3). We used regression modelling to explore whether factors such as age or the intensity of malaria transmission in the trial location could explain any of the variation observed in the proportion of late treatment failures that were symptomatic.

## Methods

### Literature Review

For pre-2009 trials we used an existing Cochrane systematic review of clinical trials of ACTs for treatment of uncomplicated malaria (3). For post-2009 data we conducted a search between 20^th^ March 2019 and 30^th^ March 2019 of PubMed, Medline and Embase using the term ‘artemether-lumefantrine’, limited to publications from 1^st^ January 2009. Details of the search strategy can be found in the Supplementary Materials. Results were limited to randomised controlled trials using the pre-set Ovid filters in each database. Search results were not restricted by language or geographical location. Grey literature, in particular conference abstracts, were also reviewed using results retrieved from this search.

### Inclusion criteria

We included clinical trials of AL for treatment of uncomplicated malaria which reported PCR-corrected treatment failure rates and whether these failures were clinical or parasitological (i.e. asymptomatic). We included only studies using the standard dosing of AL: 6 doses taken over 3 days of tablets comprising 20mg of artemether and 120mg of lumefantrine. The number of tablets per dose is determined by weight, with one, two, three, and four tablets for individuals weighing 5-15kg, 15-25kg, 25-35kg, and >35kg respectively (2). Any deviation from this regimen could lead to either enhanced or diminished drug concentrations, which could influence the time required for a recrudescent infection to become detectable by microscopy or for symptoms to return. We also required that the trial involved only participants with a confirmed *P. falciparum* infection, who sought treatment for symptoms of malaria.

We excluded studies that did not report any treatment failures (after results had been PCR corrected). We also excluded studies in which multiple drug therapies were evaluated but results not sufficiently stratified by therapy.

From these studies, data for the number of subjects enrolled, the number and type of treatment failures observed, the duration of follow up, and the location of the trial was extracted. We also extracted information on the inclusion criteria (if any were reported) for baseline parasite density in each trial. We also recorded the age range eligible for inclusion in each trial.

Titles and abstracts were reviewed against the inclusion criteria. Any discrepancies regarding inclusion/exclusion of an article was agreed upon with the consultation of fellow reviewers.

### Estimates of transmission intensity in each trial site

We used estimates of *Plasmodium falciparum* malaria prevalence by microscopy in children between 2 and 10 years of age (P*f*PR_2-10_) by the Malaria Atlas Project (MAP) at the time and location of each trial (21). We utilised the R package malariaAtlas (22), which provides prevalence estimates once the desired year, longitude, and latitude are provided. For each site location and year of trial, we used the R package to extract prevalence estimates, averaged over a 20-kilometre area. The latitude and longitude of each trial site was obtained using the website www.latlong.net (23).

### Statistical analysis

We used Bayesian regression modelling to explore the variation observed in the proportion of LTFs for which symptoms returned, which we call *ρ*. In a trial with *N_tot_* LTFs, the number of these for which symptoms return (*N_LCF_*) follows a binomial distribution *i.e. N_LCF_~Bin(N_tot_,ρ)*. We used a logistic regression framework to model *ρ*. Covariates included in the model were: the transmission intensity (x_1_) at the time and location of each trial (expressed in terms of the MAP-estimated P*f*PR_2-10_), the duration of follow up in each trial (x_2_), the age range of the cohort (x_3_), and the minimum baseline parasite density that was required for patients to be included in the original trial (x_4_). For the model containing all four variables, we wrote the regression model as

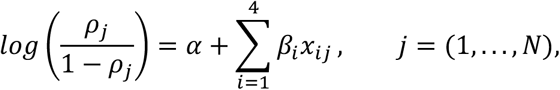

where the subscript *j* enumerates the clinical trials and *ρ* has been transformed to the logodds scale. We retained the continuous value for P*f*PR_2-10_ (expressed as a percentage) in the regression models, and constructed three categorical variables for x_2_, x_3_ and x_4_. For duration of follow up, the categorical variable was set to 1 for trials with a follow up greater than 28 days and set to 0 otherwise. For age of participants, we were particularly interested in studies that only included young children, so we set x_3_ equal to 1 for trials that enrolled participants with a maximum age of 72 months or less, and equal to 0 for all other trials. For baseline parasite density, we used a categorical variable equal to 1 for trials that allowed individuals with a baseline parasitaemia of ≥2×10^5^ parasites per microlitre to be enrolled, and equal to 0 otherwise.

The regression models were fitted in R using RStan (24), via the rethinking package (25). As we did not have any information from previous studies to inform the parameter values, we did not incorporate any prior information into the fitting procedure, although we used weakly informative priors to ensure regularisation. The goodness of fit of candidate models, containing all or some of these variables, were compared using the Watanabe Akaike Information Criterion (WAIC), which penalises each model according to the number of parameters included (26).

## Results

The 2009 Cochrane review, ‘Artemisinin-based combination therapy for treating uncomplicated malaria’ (24) identified 18 studies meeting the inclusion criteria for our study. Our search strategy for studies published after 2009 returned 643 results after removing duplicates (Figure 1). Titles and abstracts were screened against the inclusion and exclusion criteria, with full texts reviewed for 335 studies. A high number of studies (84) that would otherwise have met the inclusion criteria could not be included, as LTFs were not stratified into LCFs and LPFs. Of the 60 studies included in the analysis, several were multi-centred trials that reported results separately for each site, resulting in 75 measures of the proportion of LTFs which were symptomatic.

**Figure 1:**
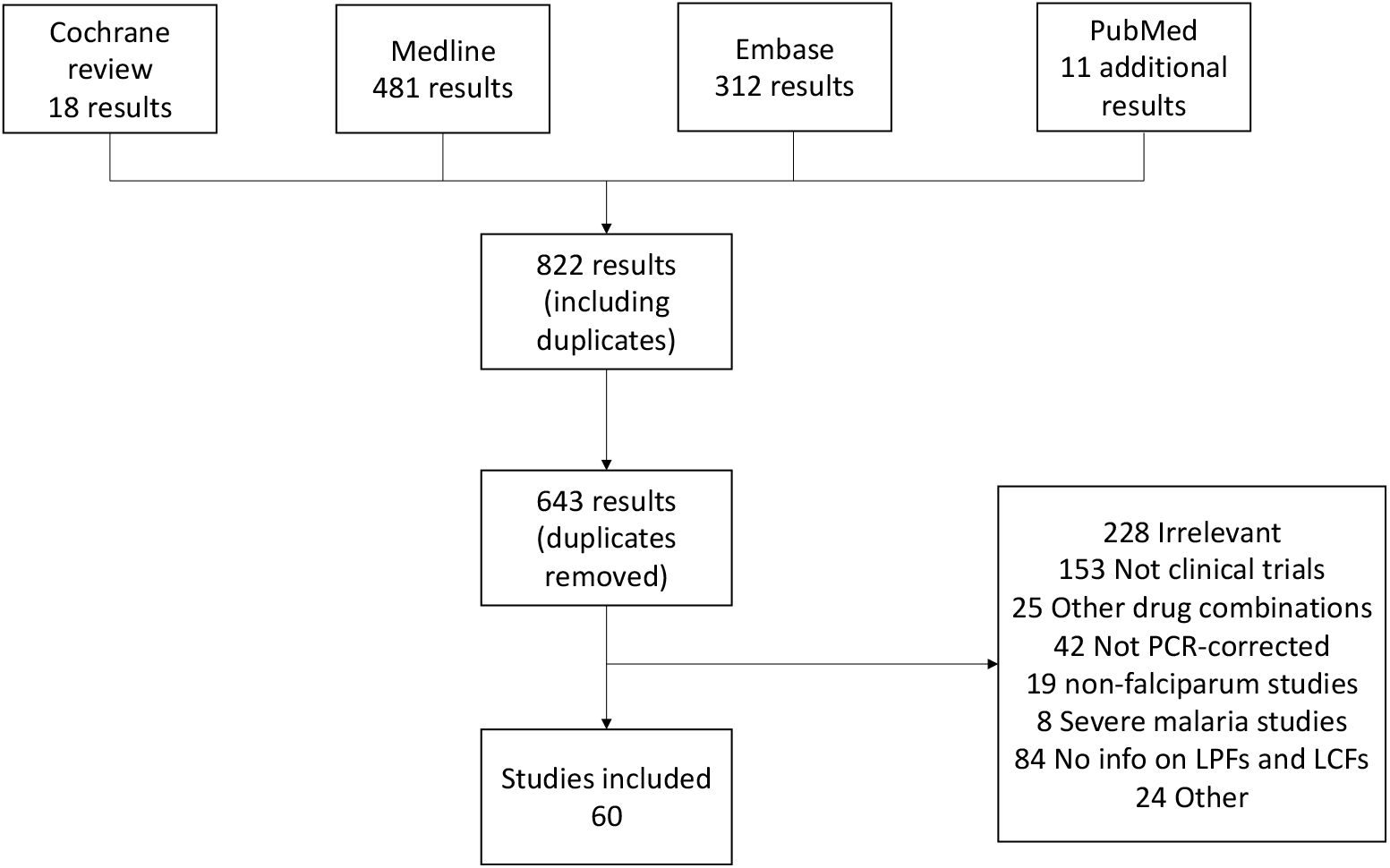
Flowchart showing the origin of all studies included in this review.

Across the included studies, 14 ETFs and 397 LTFs (after PCR-correction) were recorded out of 9137 patients who were enrolled and remained in the trial until at least the first day of follow-up. This results in an overall failure rate of 4.5% (95% CI 4.1-4.9%) for this population. Of the late treatment failures, 162 were recorded as late clinical failures (40.8%, 95% CIs [35.9-45.8%]). The majority of studies (81 %) reported PCR-corrected results for a single follow-up day, while none had more than two (19%). The most common time point (81% of studies) for reporting results was 28 days after treatment commenced, although a substantial number (33%) reported results at day 42. A number of trials enrolled only young children, especially in high transmission areas. For example, 37% of the trials included here had a maximum age for enrolment of 72 months or less. Of the studies included, there were no trials that only enrolled adults. The transmission intensity at the trial location, as estimated from the Malaria Atlas Project, varied widely with slide prevalence in 2-10 year olds ranging from 0.2% to 86.5%. Not all studies reported information on whether they used patient inclusion criteria based on parasite densities upon presentation. For those that did, the most frequent minimum parasite density required for inclusion in a trial were 1000 (48% of trials) and 2000 (35% of trials) parasites per microlitre. For the maximum parasite density, 17% of trials set a value of 10^5^ parasites per microlitre, whilst 53% allows individuals with a baseline parasitaemia of up to 2×10^5^ parasites per microlitre to be enrolled. One study permitted patients with a baseline parasite density of up to 5×10^5^ parasites per microlitre to be enrolled.

### Factors affecting the proportion of symptomatic treatment failures

We explored whether any of the variation in the proportion of late treatment failures that were recorded as LCFs could be explained by transmission intensity (represented by the estimate of malaria prevalence by microscopy in children aged 2-10, P*f*PR_2-10_), duration of follow up, age range of the cohort, or whether patients with very high baseline parasitaemia were enrolled in the study (Table 1). Of the 75 trials summarised in the previous subsection, we dropped 5 from this analysis because they were carried out across multiple countries and we were unable to fully stratify the results by location. Using data from the remaining 70 trials, we constructed logistic regression models (see Methods for full details).

**Table 1:** Summary of trial data used for regression modelling. These data come from 60 published studies, splitting multi-centred studies by site (see full data in SI). This table does not include data from 5 multi-country studies for which the data could not be stratified by site (these data are included in the findings reported in the Results section).

| Quantity | Value |
| --- | --- |
| Total number of trials | 70 |
| Reporting days | 2 trials reported on Day 14, 56 trials on Day 28, 22 trials on Day 42, one trial on Day 45, one on Day 56 |
| MAP prevalence estimates (mean and range) | 25.9% [0.2%,86.5%] |
| Trials treating young children only (<72 months) | 26 |
| Total number of patients treated | 8616 |
| Patients treated in each trial (mean and range) | 123 [23,665] |
| Total number of patients for whom treatment failed | 14 ETFs, 380 LTFs (of which 219 were late parasitological failures and 161 [42.4%] were late clinical failures) |

We assessed the goodness of fit for each model (containing combinations of these four variables), with a penalty for the number of parameters included. The leading 5 candidate models, ordered by goodness of fit, are summarised in Table 2. The best fit model contained two of the four covariates, those for transmission intensity and age (Figure 2). We find that a lower proportion of treatment failures are symptomatic in settings with a high malaria prevalence. The model predicts a 1.17-fold reduction (95% CIs, 1.08-1.26) in the odds of symptoms returning in recrudescent infections for every 10% increase in prevalence. In this model, trials that only enrolled young children are expected to have a higher proportion of symptomatic treatment failures, increasing the odds of symptoms returning by 1.61-fold (95% CIs, 1.01-2.59). We illustrate our findings with an example: in a trial that only enrolled young children, the model predicts that 60.9% (95% CIs, 46.9-72.4%) of late treatment failures would be symptomatic if carried out in a location with a P*f*PR_2-10_ of 10%, compared with 45.8% (95% CIs, 36.5-54.5%) if the same trial was carried out in a location with a P*f*PR_2-10_ of 50%.

**Table 2:**
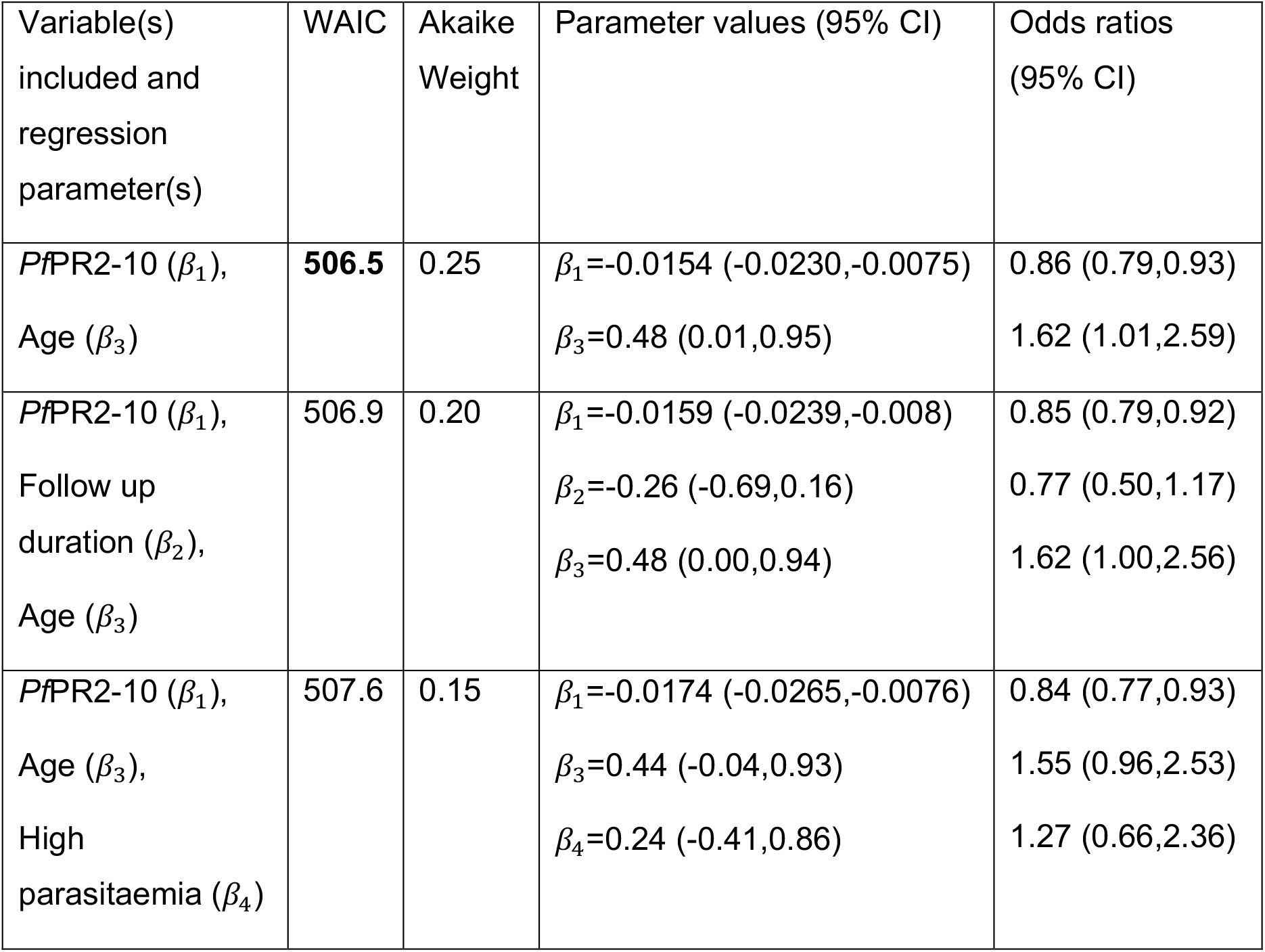
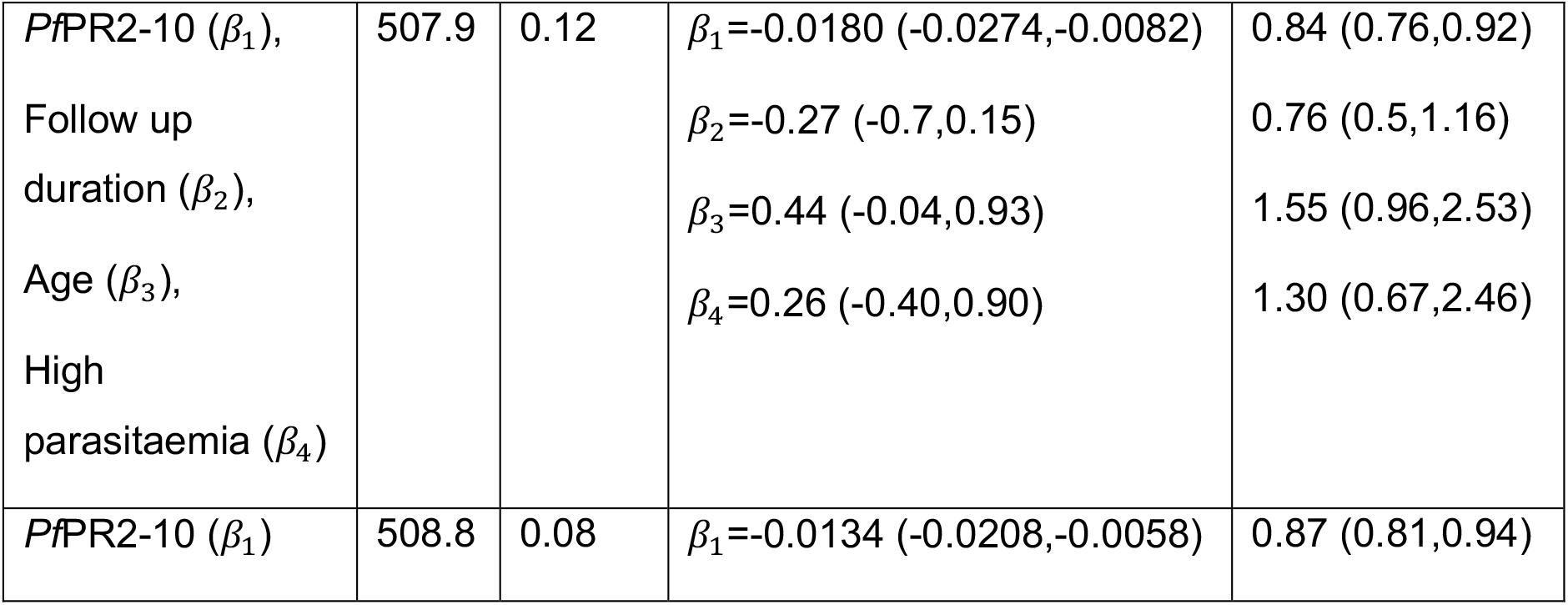
Factors associated with the proportion of late treatment failures that were symptomatic. We compared the fit of a number of uni- and multivariate regression models. The variables included were: the MAP-estimated malaria prevalence in children 2-10 years of age at the time and location of each trial (*Pf*PR_2-10_, expressed as a percentage); the duration of follow up in each trial (categorical variable set to 1 if the follow-up period exceeded 28 days); age range of the cohort enrolled (categorical variable set to 1 if age of participants restricted to ≤ 72 months); high-density infections permitted in the trial (categorical variable set to 1 if baseline parasitaemia ≥2× 10^5^ parasites per microlitre eligible for inclusion). The model fit was assessed by WAIC. The model that included variables for malaria prevalence and the age range of the cohort provided the best fit to the data. We also list some of the other candidate models, ordered by WAIC, and the Akaike weight associated with each model in the ensemble. Results generated from the best-fit model are shown in Figure 2. The results shown in Supplementary Figure 1 were generated from the ensemble of models presented here, using the models’ Akaike weights. Of the univariate models, only the model that included *Pf*PR_2-10_ outperformed the intercept-only model (WAIC score 520.1). Parameter values and corresponding odds ratios are summarised by the posterior means and the 95% credible intervals. Note that for *Pf*PR_2-10_, the ORs shown correspond to an increase in prevalence of 10%.

**Figure 2:**
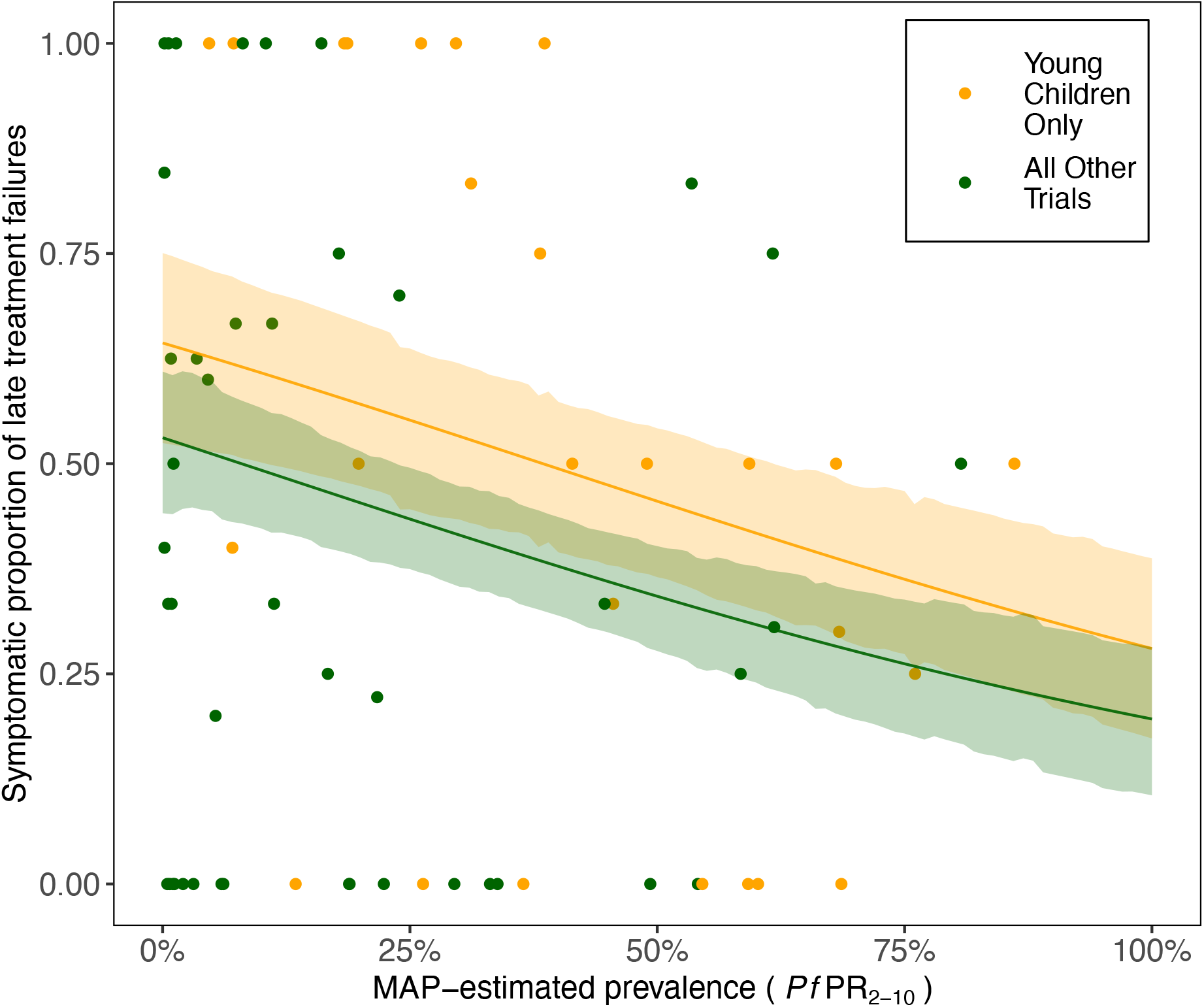
Model predictions for the proportion of late treatment failures that are symptomatic. The proportion of treatment failures that are symptomatic decreases with transmission intensity, and is also lower in young children. Points show data, and lines are predictions from the multivariable regression modelling with 95% credible intervals as shaded areas. Results were generated from the best-fit model (Table 2), which included covariates for transmission intensity (measured as malaria prevalence) and age (binary variable, indicating trials that only enrolled children under 72 months old).

We conclude this section by noting that the best-fit model only narrowly out-performed some of the other candidates (Table 2). As a sensitivity analysis, we also generated output from an ensemble of regression models, weighted by their goodness of fit (Supplementary Figure 1) (26). The generated model predictions are very similar to those shown in Figure 2, with slightly wider credible intervals (see caption of Supplementary Figure 1, where we repeat the worked example carried out in the previous paragraph).

## Discussion

In this work we have carried out a systematic review of clinical trials in which AL was used to treat uncomplicated *Plasmodium falciparum* malaria. Among individuals whose infections recrudesced following treatment, we found that 40.8% (95% CIs [35.9-45.8%]) of late treatment failures were classified as LCFs, treatment failures which experience the return of clinical symptoms. We found associations between a higher proportion of treatment failures being symptomatic and younger age as well as lower transmission settings, consistent with lower prior exposure and immunity in these study populations. The number of treatment failures observed in an individual trial is usually very low (here, the median number was 3, range [1,72]), which means that the results presented here could only be obtained via a pooled analysis.

The lack of recurrent symptoms in the majority of patients when they fail treatment suggests they would be unlikely to seek further treatment despite having persistent parasitaemia. These parasites have passed through drug pressure, giving any parasites with newly emerging or spreading drug resistant mutations a survival advantage. We do not know how long these recurrent infections would last, but untreated infections are often chronic and last on average about 6 months (27), potentially conferring a large transmission advantage on surviving parasites. Such a process might be particularly important during the early stages of drug-resistance evolution. Malaria parasites often acquire drug-resistance mutations in a step-wise fashion (28), with a first mutation conferring only a low level of resistance, followed by additional mutation conferring greater resistance (although few parasites are ever 100% resistant to a drug). Now that combination therapies are the norm for malaria treatment, the parasites must also acquire resistance to both drugs. When a parasite is only partially resistant, a second course of treatment in patients with recurrent symptomatic treatment may be enough to clear all the parasites (particularly if a different drug is given, e.g. an ACT with a different partner drug). Therefore, understanding the probability of re-treatment after initial treatment failure is important to quantifying the potential spread of resistance, for example in mathematical models (29).

One interesting avenue for further work would be to repeat the analysis for another ACT. In particular, it would be interesting to see if different results were obtained for dihydroartemisinin-piperaquine, as piperaquine has a much longer half-life than lumefantrine (30), and might be expected to suppress recrudescent infections for a longer period of time thereby delaying the reappearance of symptoms. To understand the role of recurrent symptoms in the spread of drug resistance, it would also be useful to collect similar data from trials where drug resistance is present.

One limitation of using this approach to estimate the proportion of treatment failures that become symptomatic is that it is restricted by the duration of follow-up in each trial. We cannot be sure if a patient would become symptomatic beyond the end of the follow-up period. Equally, an asymptomatic infection detected during follow-up and treated within the trial setting may have later developed into a symptomatic infection in a non-trial setting. In the regression modelling, we did not find a strong effect of the duration of follow up on the likelihood of symptoms returning after treatment failed. However, the dataset has very little temporal resolution (83% of studies had only reported results for one time point), which made it difficult to scrutinise this in any detail. Outside trial settings, AL failure rates are likely to be somewhat higher due to imperfect adherence to treatment (31,32). It is possible that the likelihood of symptoms returning upon recrudescence of parasitaemia would be different in patients who fail due to lower adherence.

Not all studies provided information on the inclusion criteria for baseline parasite density in each trial. For studies that did not report it, we assumed that any individual with uncomplicated symptoms who was positive for parasites by microscopy could be enrolled in the trial. Furthermore, the connection between the range of parasitaemia permitted in each trial and the distribution of parasitaemia measured in the cohort is not a clear one, but this was not available for all trials either.

In recent within-host modelling work (15,31) we investigated the link between poor adherence and treatment failure, with the more recent study translating treatment failure into an enhanced capacity to transmit malaria. It was shown that the proportion of recrudescent infections that are retreated had a big impact on onward transmission (15), but little insight was provided as to what, in a particular setting, this proportion would be. In reality, this will depend on many factors, not least the availability and affordability of a second course of treatment. However, outside of a trial setting it is very unlikely that an asymptomatic recrudescent infection would be detected and retreated.

## Conclusion

By pooling together data from a large number of clinical trials, we were able to gain insight into the likelihood of patients becoming symptomatic again following treatment failure. We show that this is more likely in younger children and in lower transmission settings. Asymptomatic treatment failure could help drug resistant parasites to spread in high transmission settings.

## Data Availability

All data and code will be available upon reasonable request after publication

**List of Abbreviations**
ACT: artemisinin-based combination therapy
AL: artemether-lumefantrine
PCR: Polymerase chain reaction
ETF: early treatment failure
LTF: late treatment failure
LCF: late clinical failure
LPF: late parasitological failure
MAP: Malaria Atlas Project
P*f*PR_2-10_: *Plasmodium falciparum* prevalence in children aged 2-10 years
WAIC: Watanabe-Akaike Information Criterion
OR: odds ratio

## Ethics and consent for participation

Not applicable.

## Consent for publication

Not applicable.

## Availability of Data and Materials

All data and code will be made available upon reasonable request from the corresponding author.

## Authors’ Contributions

JDC and LCO designed the study. RM carried out the systematic review. JDC and RM analysed the data, with all authors contributing to the interpretation of the findings. JDC wrote the first draft of the manuscript. All authors contributed to the manuscript and approved the final version.

## Acknowledgements

JDC would like to thank Nathan Green for useful discussions.

## References

1. World Health Organization. World Malaria Report 2019. Geneva; 2019.

2. World Health Organization. Guidelines for the Treatment of Malaria. 2015., 3rd edition. Geneva: 2015.

3. Sinclair D, Zani B, Donegan S, Olliaro P, Garner P. Artemisinin-based combination therapy for treating uncomplicated malaria. Cochrane Database of Systematic Reviews. 2009.

4. Ezzet F, Van Vugt M, Nosten F, Looareesuwan S, White NJ. Pharmacokinetics and pharmacodynamics of lumefantrine (benflumetol) in acute falciparum malaria. Antimicrob Agents Chemother. 2000;44(3):697–704.

5. WorldWide Antimalarial Resistance Network Lumefantrine PKPD Study Group. Artemether-lumefantrine treatment of uncomplicated Plasmodium falciparum malaria: a systematic review and meta-analysis of day 7 lumefantrine concentrations and therapeutic response using individual patient data. BMC Med. 2015;13:227.

6. Kloprogge F, Workman L, Borrmann S, Tekete M, Lefevre G, Hamed K, et al. Artemether-lumefantrine dosing for malaria treatment in young children and pregnant women: A pharmacokinetic-pharmacodynamic meta-analysis. PLoS Med. 2018;15(6):e1002579.

7. Creek DJ, Bigira V, McCormack S, Arinaitwe E, Wanzira H, Kakuru A, et al. Pharmacokinetic predictors for recurrent malaria after dihydroartemisinin-piperaquine treatment of uncomplicated malaria in ugandan infants. J Infect Dis. 2013;207(11):1646–1654.

8. Tarning J, Zongo I, Somé FA, Rouamba N, Parikh S, Rosenthal PJ, et al. Population pharmacokinetics and pharmacodynamics of piperaquine in children with uncomplicated falciparum malaria. Clin Pharmacol Ther. 2012;91(3):497–505.

9. Chotsiri P, Denoeud-Ndam L, Baudin E, Guindo O, Diawara H, Attaher O, et al. Severe Acute Malnutrition Results in Lower Lumefantrine Exposure in Children Treated With Artemether-Lumefantrine for Uncomplicated Malaria. Clin Pharmacol Ther. 2019;106(6):1299–309.

10. Staehli Hodel EM, Csajka C, Ariey F, Guidi M, Kabanywanyi AM, Duong S, et al. Effect of Single Nucleotide Polymorphisms in Cytochrome P450 Isoenzyme and N-Acetyltransferase 2 Genes on the Metabolism of Artemisinin-Based Combination Therapies in Malaria Patients from Cambodia and Tanzania. Antimicrob Agents Chemother. 2013;57(2):950–8.

11. Elewa H, Wilby KJ. A Review of Pharmacogenetics of Antimalarials and Associated Clinical Implications. Eur J Drug Metab Pharmacokinet. 2017;42(5):745–56.

12. Bruxvoort K, Goodman C, Patrick Kachur S, Schellenberg D. How patients take malaria treatment: A systematic review of the literature on adherence to antimalarial drugs. PLoS One. 2014;9(1).

13. Banek K, Lalani M, Staedke SG, Chandramohan D. Adherence to artemisinin-based combination therapy for the treatment of malaria: A systematic review of the evidence. Malar J. 2014;13:7.

14. Ashley EA, Stepniewska K, Lindegårdh N, Annerberg A, Kham A, Brockman A, et al. How much fat is necessary to optimize lumefantrine oral bioavailability? Trop Med Int Heal. 2007;12(2):195–200.

15. Challenger JD, Gonçalves BP, Bradley J, Bruxvoort K, Tiono AB, Drakeley C, et al. How delayed and non-adherent treatment contribute to onward transmission of malaria: A modelling study. BMJ Glob Heal. 2019;4:e001856.

16. Bretscher MT, Maire N, Felger I, Owusu-Agyei S, Smith T. Asymptomatic Plasmodium falciparum infections may not be shortened by acquired immunity. Malar J. 2015; 14(1).

17. Bretscher MT, Maire N, Chitnis N, Felger I, Owusu-Agyei S, Smith T. The distribution of Plasmodium falciparum infection durations. Epidemics. 2011;3(2):109–18.

18. World Health Organization. Methods for surveillance of antimalarial drug efficacy. Geneva; 2009.

19. Atwine D, Balikagala B, Bassat Q, Chalwe V, D’Alessandro U, Dhorda M, et al. A Head-to-Head Comparison of Four Artemisinin-Based Combinations for Treating Uncomplicated Malaria in African Children: A Randomized Trial. PLOS Med. 2011 Nov 1;8(11).

20. World Health Organization. World malaria report 2018. Geneva; 2018.

21. Weiss DJ, Lucas TCD, Nguyen M, Nandi AK, Bisanzio D, Battle KE, et al. Mapping the global prevalence, incidence, and mortality of Plasmodium falciparum, 2000–17: a spatial and temporal modelling study. Lancet. 2019;394:322–31.

22. Pfeffer D, Lucas T, May D, Harris J, Rozier J, Twohig K, et al. malariaAtlas: an R interface to global malariometric data hosted by the Malaria Atlas Project. Malar J. 2018;17(1):352.

23. www.latlong.net [Internet]. Available from: www.latlong.net

24. Team SD. RStan: the R interface to Stan. 2020. p. R package version 2.21.1.

25. McElreath R. rethinking: Statistical rethinking book package. 2020. p. R package version 2.01.

26. McElreath R. Statistical rethinking: A bayesian course with examples in R and stan. 1st ed. Statistical Rethinking: A Bayesian Course with Examples in R and Stan. Chapman and Hall/CRC; 2016.

27. Felger I, Maire M, Bretscher MT, Falk N, Tiaden A, Sama W, et al. The Dynamics of Natural Plasmodium falciparum Infections. PLoS One. 2012;7(9).

28. Lozovsky ER, Chookajorn T, Brown KM, Imwong M, Shaw PJ, Kamchonwongpaisan S, et al. Stepwise acquisition of pyrimethamine resistance in the malaria parasite. Proc Natl Acad Sci U S A. 2009;106(29):12025–30.

29. White NJ, Pongtavornpinyo W, Maude RJ. Hyperparasitaemia and low dosing are an important source of anti-malarial drug resistance. Malar J. 2009;8:253.

30. Okell LC, Cairns M, Griffin JT, Ferguson NM, Tarning J, Jagoe G, et al. Contrasting benefits of different artemisinin combination therapies as first-line malaria treatments using model-based cost-effectiveness analysis. Nat Commun. 2014;5.

31. Challenger JD, Bruxvoort K, Ghani AC, Okell LC. Assessing the impact of imperfect adherence to artemether-lumefantrine on malaria treatment outcomes using within-host modelling. Nat Commun. 2017;8(1):1373.

32. Schoepflin S, Lin E, Kiniboro B, DaRe JT, Mehlotra RK, Zimmerman PA, et al. Treatment with coartem (artemether-lumefantrine) in Papua New Guinea. Am J Trop Med Hyg. 2010;82(4):529–34.

